# Scoping review of the evidence concerning the unique needs and experiences of Orthodox Jewish couples using maternity services

**DOI:** 10.1101/2025.10.12.25337836

**Authors:** Michal Rosie Meroz, Carol Rivas, Christine McCourt

## Abstract

**Background:** of the 300,000 Jews living in the UK today, 19% identify as Orthodox. High fertility rates within some of the community sub-groups make interactions with NHS maternity services frequent. The unique lifestyle and religious customs of community members have the potential to make these interactions complex and confusing for both NHS staff and community members.

**Aims:** to identify relevant studies and map the literature concerning the interaction of Orthodox Jewish couples with maternity services and identify gaps in the literature.

**Method:** a scoping review, undertaken in line with the methodological framework of Arksey and O’Malley, following PRISMA and JBI guidelines. We searched CINAHL, SocINDEX, Medline, ProQuest, Web of Science and grey literature.

**Results:** of the twelve included studies, ten were qualitative and two were quantitative. The geographical locations included Israel and North America, with no studies conducted in the UK. Half of the studies were carried out over 10 years ago. Using Thomas and Harden’s thematic synthesis, five analytical themes were identified: 1. Between the Divine and the Earthly: Negotiating Faith and Modern Medicine. 2. Holding Space or Holding Back: The Role of the Husband and the Community. 3. Born to Become a Mother, through Joy and Hardship. 4. Praying Quietly: Coping Mechanisms. 5. Keeping it Kosher: Religious and Maternity Care.

**Conclusion:** the maternity experiences of Orthodox Jewish couples reflect unique needs shaped by a complex interplay of faith and risk-based medicine. The centrality of the community serves as a double-edged sword, offering vital support while also imposing restrictions and norms. There is an urgent need for high-quality research in the UK into the needs and experiences of Orthodox Jewish couples to inform NHS policies.

## Background

Judaism can be described as ethnicity, culture and religion. While most 300,000 UK Jews share the same customs and festivals, their religious affiliations span a spectrum from secular to Reform, Masorti, Central Orthodox (or modern Orthodox), and strictly Orthodox (Haredi) [1, 2].

Within this spectrum, the Haredi (Ultra-Orthodox) are the most observant and insular subgroup. An estimated fertility rate of seven children per woman [3–5], compared to 1.44 in the UK average in 2023 [6], makes them one of the fastest-growing communities in the world. Following Israel and North America, the UK has the third-largest Haredi community in the world, with approximately 76000 members (named “Haredim”) [3]. Despite the distinction between Orthodox and Haredi, these terms are sometimes used interchangeably, and defining oneself as a “Haredi” is based on individual perception [7]. Therefore, we will use the term “Orthodox Jews” throughout this paper to refer to all Orthodox Jewish people, including Haredim, unless referring specifically to one group or another.

Orthodox Jews (OJ) follow the Halacha, a set of ancient Jewish laws that regulate all aspects of life, from the cradle to the grave, including dress code, dietary restrictions, public behaviour, religious rituals, and Shabbat (Saturday, the holy day of rest) and holy days observance [8–10]. The Halacha, and rabbinical authorities interpreting it, help guide OJ families through decision-making around sexuality, the use of contraception, screening tests in pregnancy, birth interventions and care for the mother and her newborn [9, 11–13]. Halachic laws also determine the level of involvement and how the husband is expected to support his wife during childbirth, while traditionally avoiding any physical contact [11].

With origins dating back approximately 3000 years ago, the Halacha continues to evolve and adjust to meet the needs of the modern world [8, 9, 14]. Nonetheless, certain Haredi communities around the globe remain intentionally insular, separating themselves from broader society, avoiding electronic media and smartphones and limiting interactions with people outside their community [15–18]. Scholars have described this deliberate separation as a form of “immunity” [19] against the perceived negative influence of non-Orthodox values. In this context, rabbis have been described as gatekeepers, safeguarding the community from what Raucher referred to as “contamination” [18].

Most Orthodox and all Haredim people are educated in Jewish schools [1], and the community is the main source of support [15, 16, 20]. In the UK, many Haredim speak Yiddish or Hebrew as their first language, and some do not speak English at all [4, 16, 21]. An insular lifestyle has also led to minimal interaction of some Haredi women in the UK with the “external” world (outside the community), with maternity services sometimes being the only such interaction [19].

The complexity of Halachic laws, combined with cultural and language barriers, sometimes results in communication barriers, misunderstandings, judgment, and a lack of trust when interacting with NHS maternity staff, as reported in the literature [15–17, 20, 22]. It has also been reported that compared to the general population, Haredim are more likely to fully or partially decline pregnancy screening tests and caesarean sections and have lower uptake of children’s vaccination programmes [20, 23]. In the UK, Judaism is not classified as a race, and religion is not a mandatory field in the census or medical records. As a result, data collection is limited, and pregnancy outcomes specific to Orthodox Jewish women are largely unavailable [24, 25].

Papers from North America and the UK suggest that OJ families have unique maternity needs that can be unfamiliar or confusing to maternity staff [8, 12, 14, 26–28]. A recent NHS Race and Health Observatory report stated that *“current NHS communication practices do not align with the cultural and religious lifestyle of the strictly Orthodox community*” [2,p.12] and that “*upskilling of NHS staff with cultural and religious competency is needed to build better relationships, improve one-to-one conversations and communications, as well as improving outcomes for Jewish patients*” [2,p.13].

This scoping review examines the needs and experiences of OJ couples when engaging with maternity services. The sensitive nature of childbirth, combined with high fertility rates, Halachic laws, and cultural perceptions, makes these interactions frequent and complex. The first author is a senior research midwife at Homerton Hospital in the London borough of Hackney, where one of the largest Orthodox Jewish communities in the world resides. This review was conducted as part of her National Institute for Health and Care Research (NIHR) fellowship. The co-authors, both experts in health, social care, and medical sciences, advised on and reviewed drafts of this paper. Patient and Public Involvement and Engagement (PPIE) work with the OJ communities in London and Manchester, undertaken alongside this review, will hopefully lead to a Participatory Research project with Orthodox Jewish communities across the UK.

## Methods

### Scoping review

To identify relevant studies and map the literature concerning Orthodox Jewish couples and their interaction with maternity services, and identify gaps in the literature, a scoping review was chosen [29].

We followed Arksey and O’Malley’s framework [30], and before commencing the review, we identified the research question, the search keywords and databases. We identified relevant studies, collated and summarised the findings and report the results herein.

### Identifying the research question

The research question for the current scoping review was: **What is known about the needs of Orthodox women and their partners who are using maternity services?** The sub-questions:

1. What is known about the experiences of Orthodox women and their partners who are using maternity services in the UK?
2. How are these needs being met in UK maternity services?

### Identifying relevant studies

We covered a range of study designs and qualities, using a broad search, as advised by experts in scoping review methodology [30]. As a first step, the first author searched the following databases: CINAHL, SocINDEX, Medline, ProQuest, and Web of Science. We planned to search academic theses on Ethos; however, due to a major cyber-attack on the British Library at the time of writing this review, this was not possible. Alongside the database search, the first author contacted community stakeholders to request any literature they considered relevant. Additionally, the first author invited PPIE members to share any relevant materials with her. Lastly, the first author conducted a backwards citation tracking to identify additional papers.

### Selection of eligible studies

We tried keeping our search as comprehensive as possible within the time constraint of the fellowship programme (six months in total, approximately three months dedicated to the scoping review). We identified and agreed on keywords before starting the search; however, this was an iterative process, and as we immersed ourselves in the data, this was adjusted. The final search strategy is presented in Table 1.

**Table 1:**
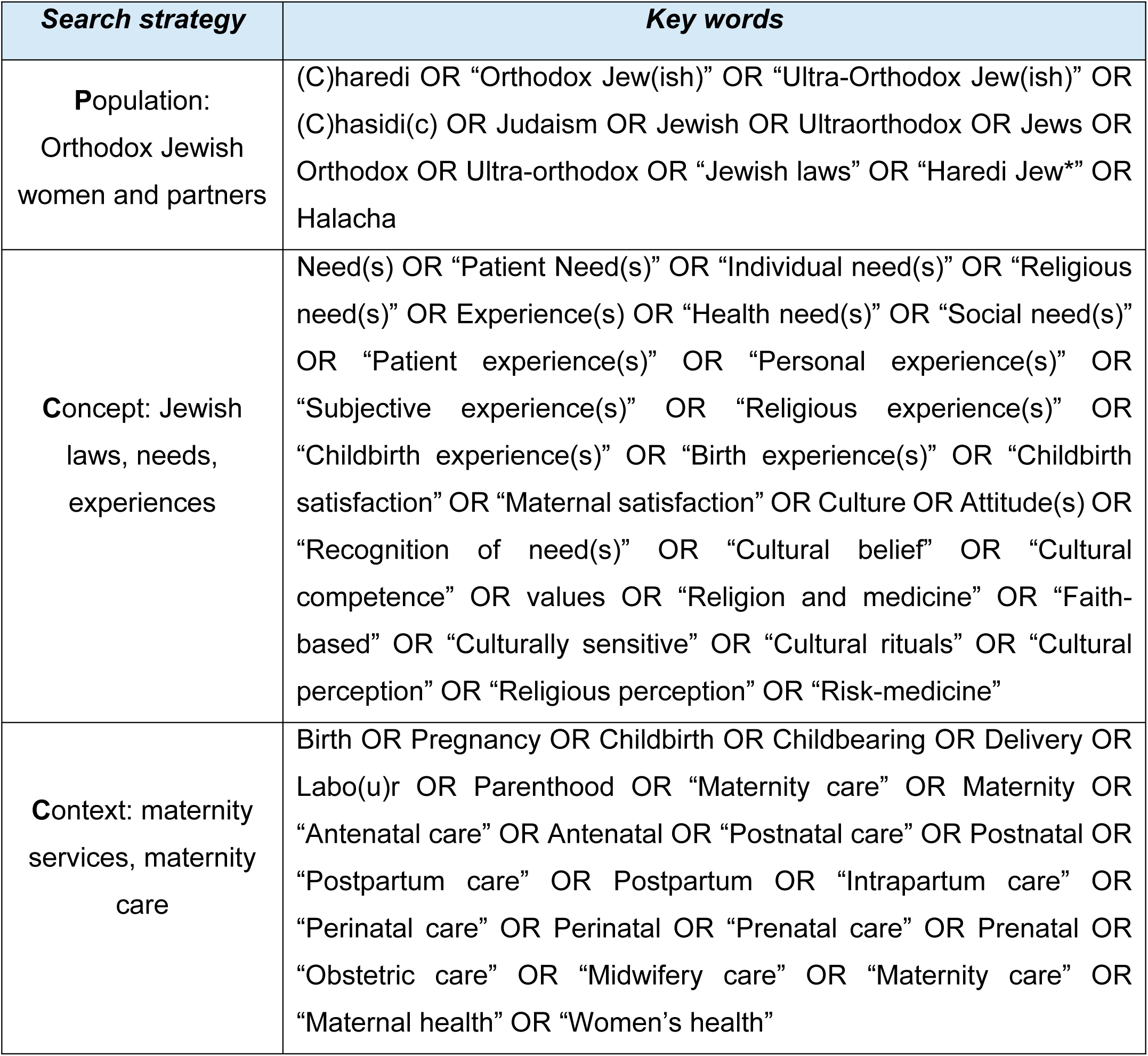
Search strategy following the PCC model [32].

The first author conducted the initial review of titles, using the above keywords. This was followed by a screening process to remove duplicates and papers that were obviously irrelevant. The first and second authors then reviewed the papers’ abstracts and full texts to identify studies that met the inclusion criteria. Disagreements were resolved by either consensus or by the third author, who acted as a third reviewer. Fig. 1 illustrates the search process, utilising the PRISMA flow diagram [31].

**Fig 1:**
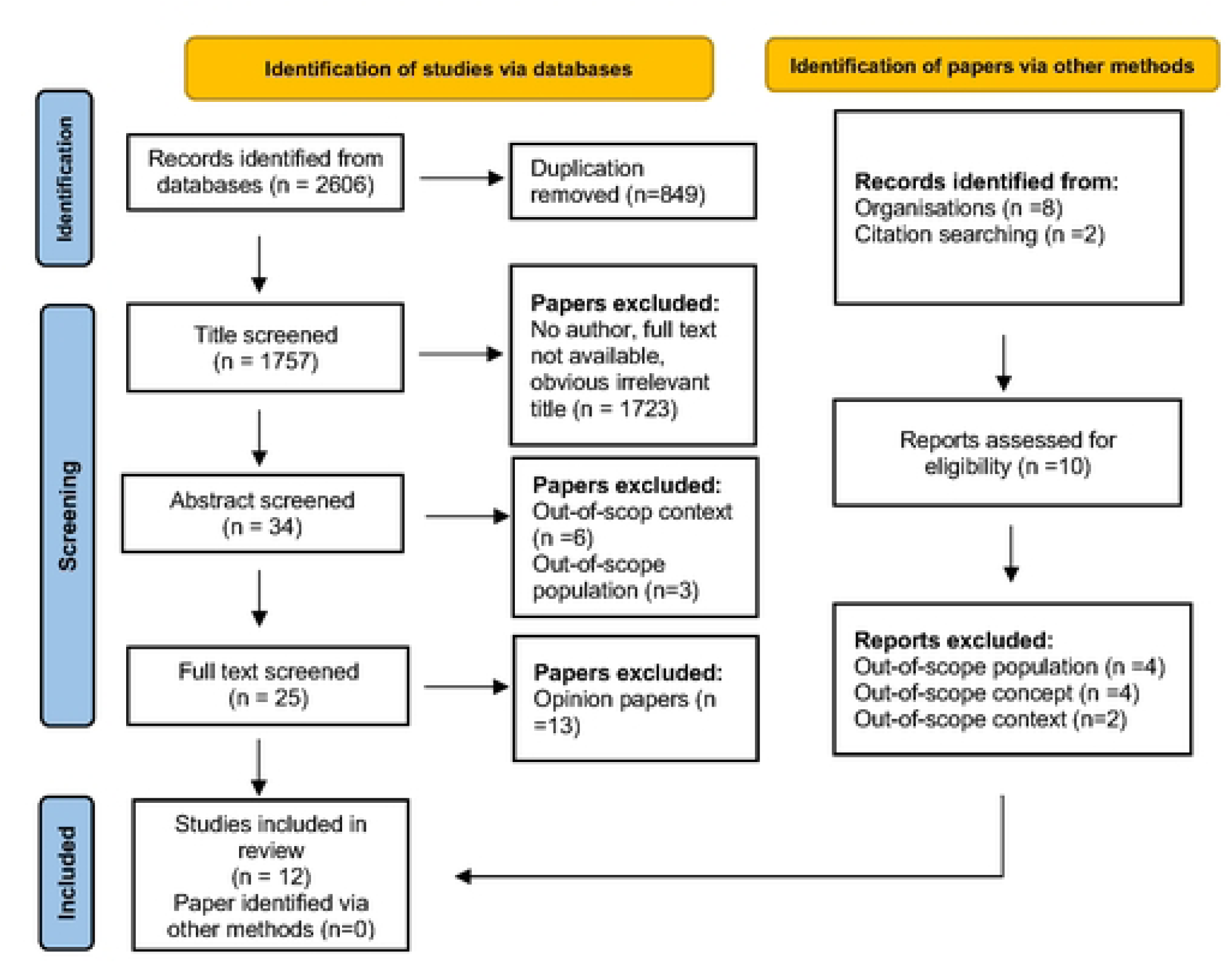
PRISMA flow diagram.

### Inclusion criteria

Following the Joanna Briggs Institute recommendations for conducting scoping reviews [32], our inclusion criteria were predefined and adhered to the Population, Concept, Context (PCC) framework as below:

**P-** Orthodox Jewish women and their partners

**C-** needs and experiences, Jewish laws

**C-** maternity services

We wanted to include original papers discussing the experiences and needs of Orthodox Jewish women and/or their partners when interacting with maternity services from conception to one year postpartum. However, due to scarce literature, we included papers that did not investigate these directly but contained data relevant to our research question. Papers that conducted interviews outside the specified postpartum timeframe were included if they addressed experiences or needs occurring within the defined period.

The paucity of relevant literature also necessitated other key decisions in our inclusion criteria. Firstly, although the Haredi is the most observant Orthodox subgroup, we chose to include all Orthodox Jews to enable us to gain an in-depth understanding of a spectrum of Orthodox experiences. Secondly, we extended the search beyond the UK to include studies from all OECD countries, to identify service gaps within UK maternity care that may be addressed differently elsewhere. We also included studies from Israel, the home to the world’s largest Orthodox Jewish population. Finally, we opted not to restrict by publication year to capture all potentially relevant studies.

### Exclusion criteria

We excluded papers with no full text or an unknown author, opinion papers and books.

### Collating and charting the data

After identifying the relevant papers, they were read independently by all authors. We then held several discussions via Microsoft Teams to explore the content of the studies. Following these discussions and consensus-building, the first author summarised the findings in an Excel spreadsheet, which was made available to all authors. As recommended by the scoping review literature, we reported the process in a transparent and detailed way to allow replication [30, 32]. This is demonstrated in Tables 1, 2.

**Table 2:**
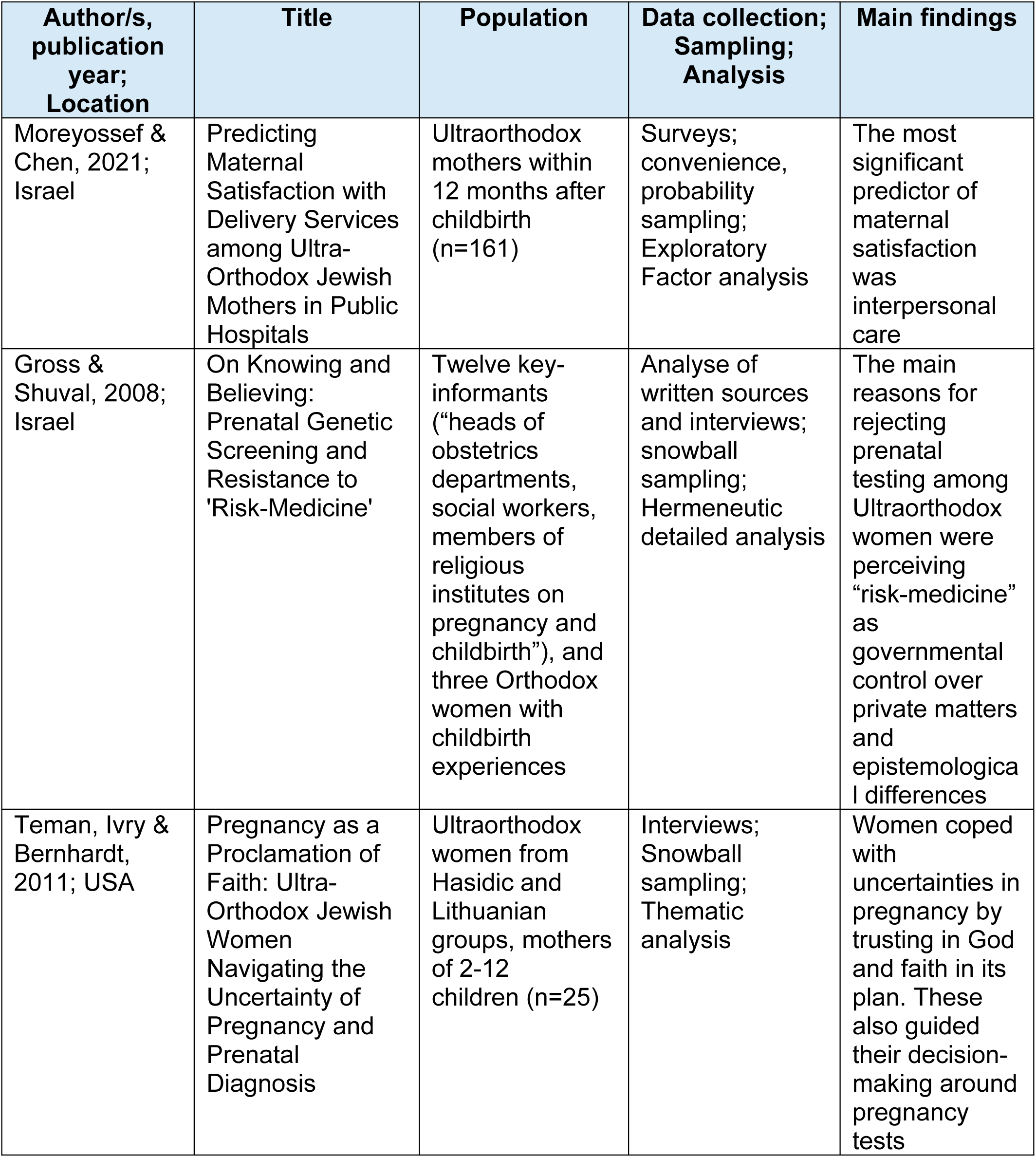
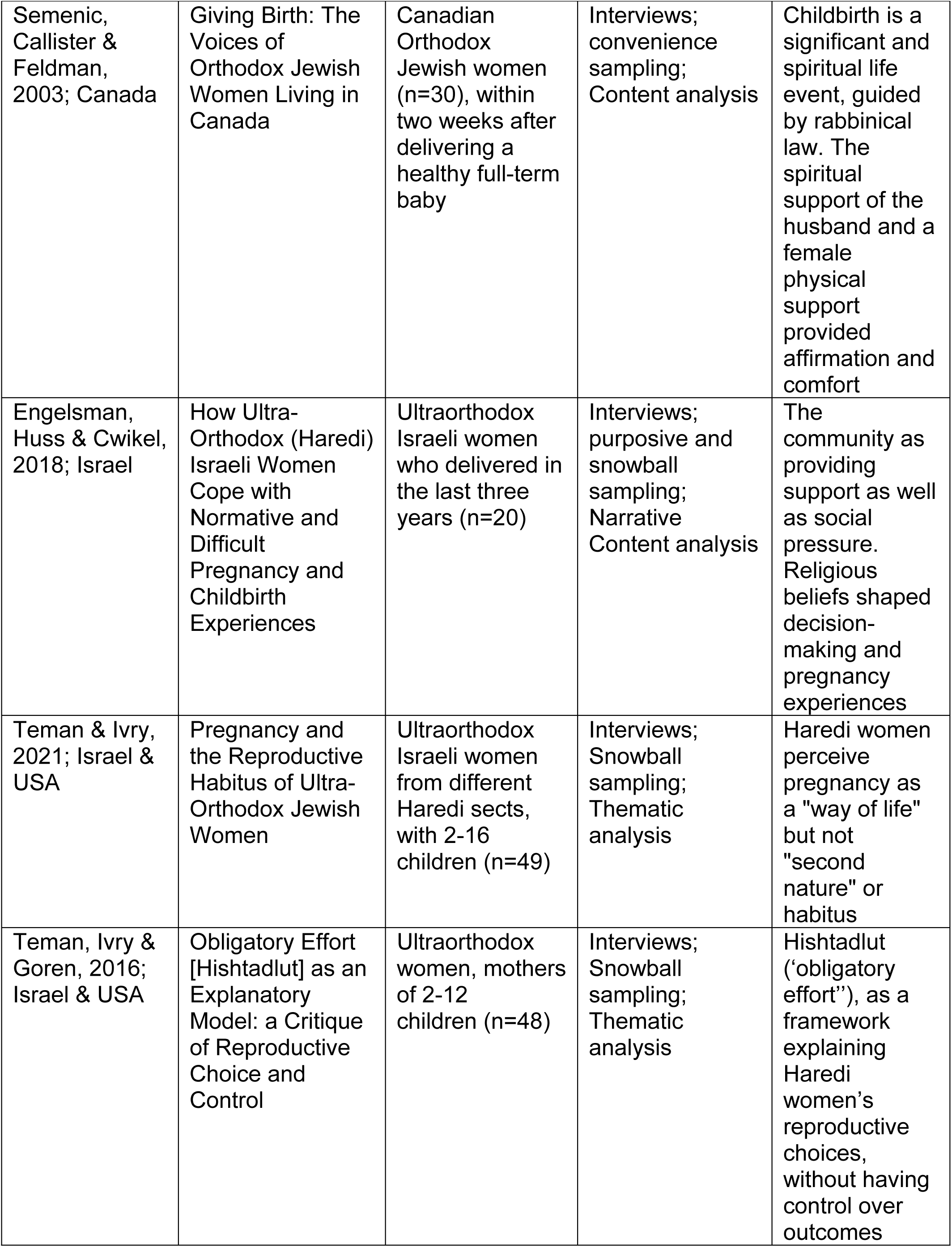
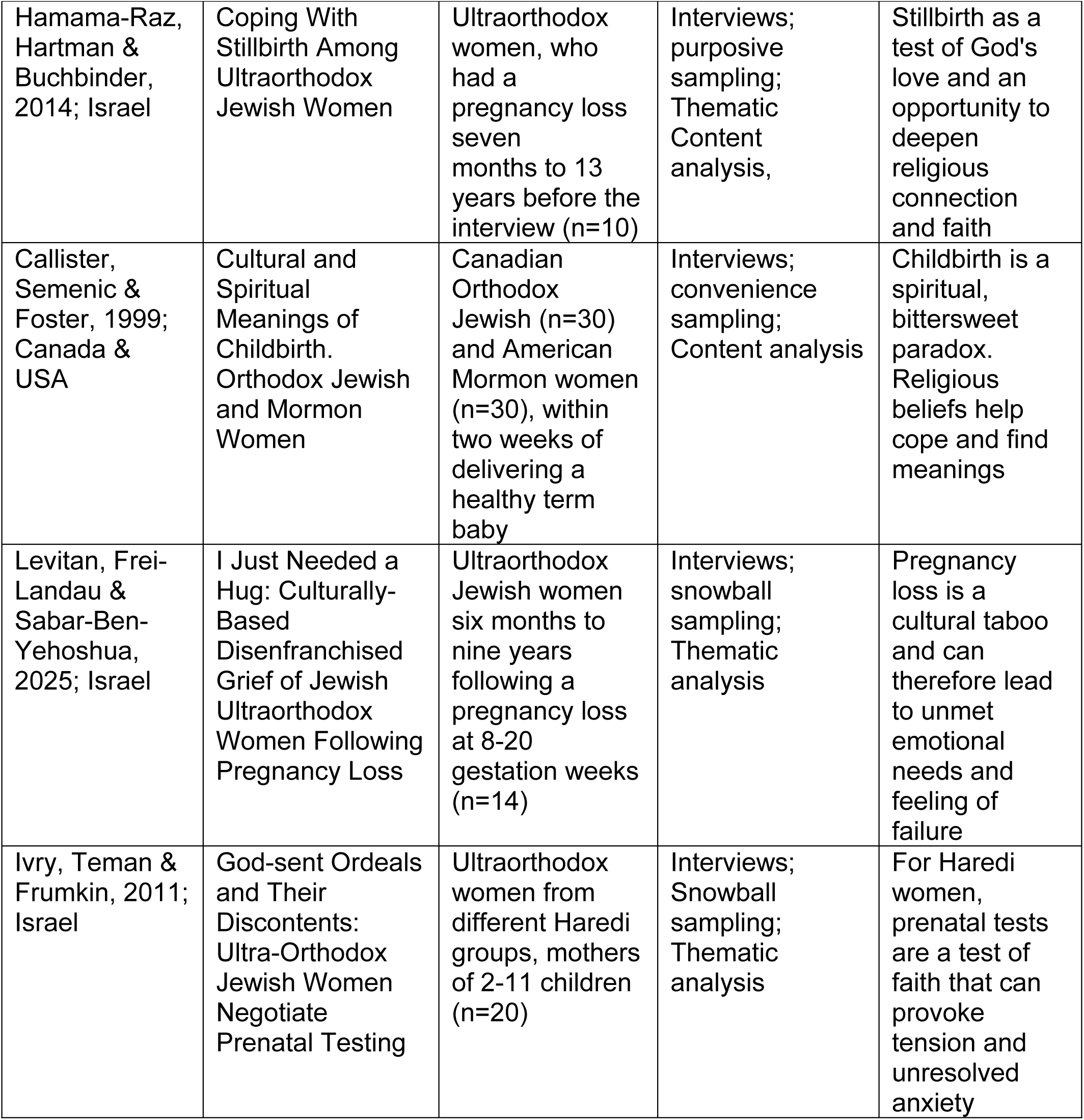
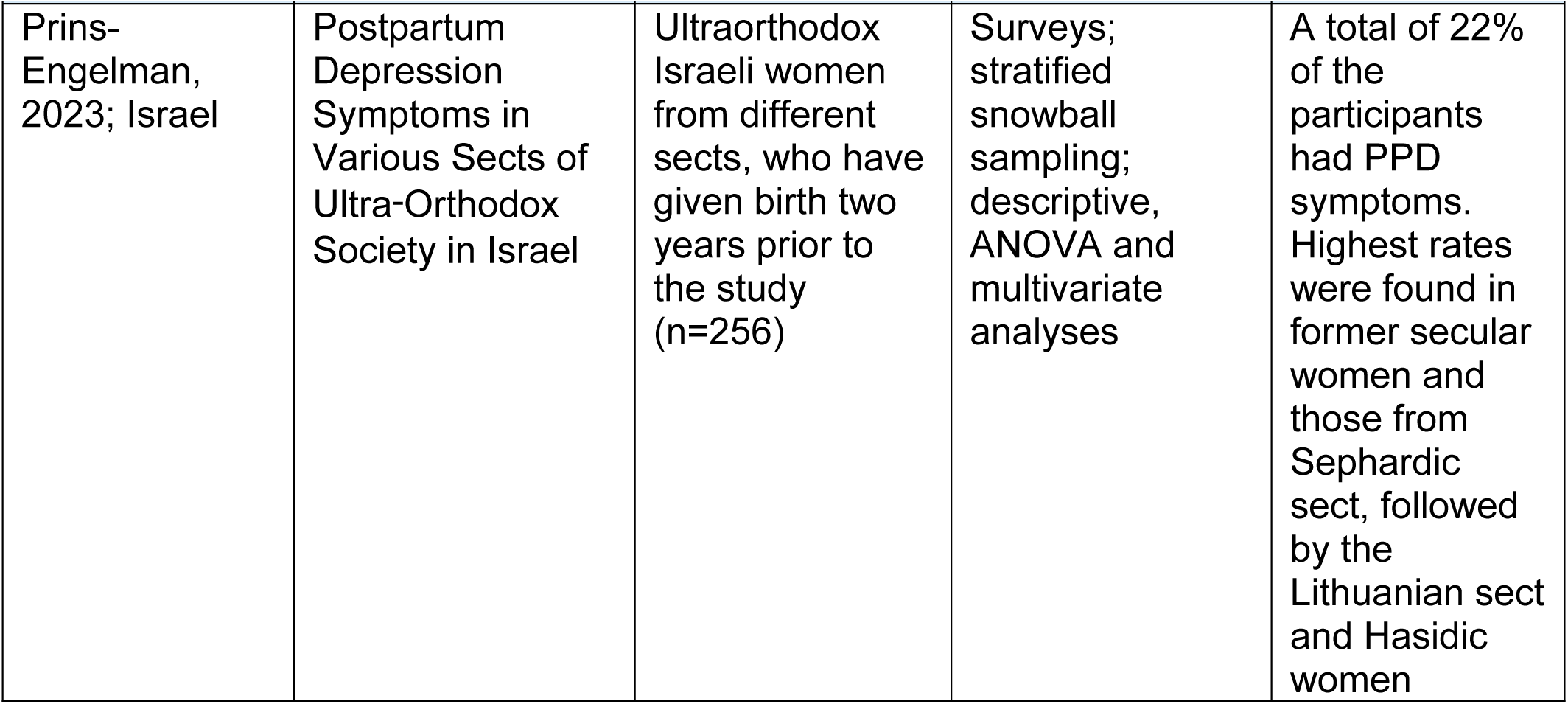
Characteristics of included studies.

## Results

### Studies characteristics

The initial database search, conducted by the first author between 8-17 April 2025, yielded 2606 papers, of which 849 were removed due to duplication. The titles of the remaining 1757 papers were screened, and 1723 were removed as they had no author, no full text available or had a title which indicated irrelevant content. Following that, the first and second authors screened the abstracts of 34 papers. Of these, three were an out-of-scope population (not Orthodox-specific) and six were of out-of-scope context (not “maternity” as defined above). The first author screened the full text for the remaining 25 papers, following which 13 were excluded as they were opinion papers with no original data. The first author conducted a backwards citation tracking of the relevant papers and identified two additional papers, which were subsequently excluded as they were out-of-scope in context. All decisions were discussed among the three authors.

Parallel to the database search, the first author emailed OJ community stakeholders and PPIE members. Eight papers were emailed, and although none of them met the inclusion criteria (one was rabbinical guidance and seven were local reports, of which only two were maternity related), we found some of them useful for our discussion.

A PRISMA flowchart describes the included and excluded studies at each stage (Fig. 1). Ten studies were qualitative, collecting data via interviews, and two studies used a quantitative design, utilising questionnaires. Seven studies were conducted in Israel, three in North America, and two in both Israel and the USA.

Of the 12 papers included in our review, one study focused on maternal satisfaction, three on genetic testing, five on the experiences and meanings of pregnancy and childbirth, and three on postpartum depression and loss (Table 2).

### Synthesis and findings

We analysed the original data presented in the Results or Findings sections of the studies. For studies that lacked clear “results” or “findings” headings, we analysed original data, typically located after the method section. We followed Thomas and Harden’s thematic synthesis process [33] and began with line-by-line coding of each study, conducted by the first author. This yielded just over 200 codes. Findings from the two quantitative studies were also coded line by line, ensuring codes accurately reflect the original results. In the next step, the codes were grouped into descriptive themes, which were then interpreted to develop analytical themes that extended beyond the original data. Some codes were linked to more than one theme. The main codes and themes are illustrated in Fig. 2, and the themes are described below.

**Fig 2:**
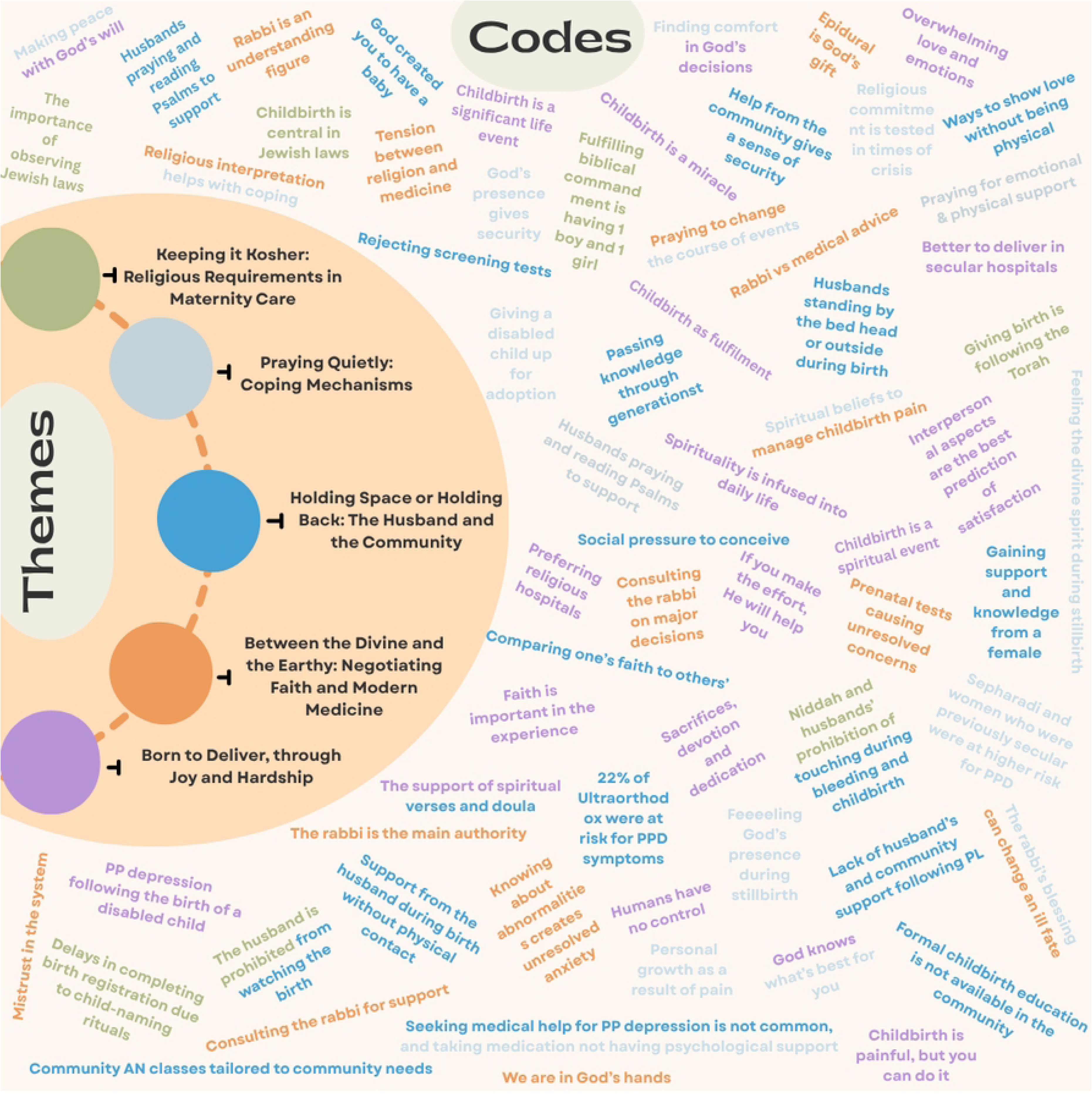
Main codes and themes.

#### **Theme One**: Between the Divine and the Earthly: Negotiating Faith and Modern Medicine

##### a. Faith in the divine will

Women described navigating maternity care decisions through a religious lens, often seeking rabbinic guidance on prenatal testing and contraception [34–39]. Most studies described the importance of the rabbi, either by direct quotes from women or indirect references. The rabbi was described as the *“Almighty’s messenger”* [36] who makes the final decision, *“the rabbi says no and that’s it…”* [40,p.274]. Engelsman and her colleagues stated that: “[women] *felt that they could express themselves with their rabbis and that the rabbis are sensitive to the needs of women”* [36,p.141].

Women described a deep faith in God as central to how they navigated uncertainty during pregnancy and childbirth. Rather than relying solely on biomedical risk assessments, many participants believed that outcomes were determined by divine force. This sense of theological certainty extended to times of loss, as one woman reflected following a stillbirth: *“we are in God’s hands. He decides who will live and who will die. I am comfortable being guided, for better or worse”* [41,p.928].

This orientation also influenced attitudes toward prenatal testing. For some women, declining tests was an expression of religious trust [37–39]. Teman and her colleagues found that in some communities, undergoing pregnancy tests could be interpreted as a sign of weak faith, undermining the belief that outcomes are in God’s hands [39]. Similarly, Ivry and her colleagues described the “ideal woman of faith” as one who surrenders to divine will, believing that if God assigns her a challenge, He will also grant her the strength to endure it [38]. The community was described as having a righteousness hierarchy, where a “*truly pious woman would not consult a rabbi: under no circumstances would she terminate* [her pregnancy]*; instead, she gracefully accepted her God-given fate”* [38,p.1532]. In other studies, some women preferred not to know to preserve the possibility of divine intervention, as one woman cited an ancient Jewish verse: *“a blessing is found only in what is hidden from the eye”* [40,p.280].

Some women described their frustration when an abnormality was found on a scan, stating that this information only creates stress and anxiety, as the Halacha does not generally permit abortions. One woman recalled her discussion with her doctor following such a diagnosis: “*why are you telling me this (when) you know I cannot do anything with this informatio*n?” [38,p.1529].

Women referred to errors in medical care as evidence that human beings lack control, reinforcing their belief in divine will [36, 38]. One woman described feeling judged by secular society for declining prenatal screening: *“The hospital gave me a feeling that I was a criminal…”* [37,p.558]. This tension was further noted by a social worker who said: “*There is something about secularity, and surely in the medical system, that is very angry at all those mothers who do not have abortions”* [37,p.558].

##### b. Biomedicine in the service of the Halacha

Although some women expressed mistrust or hesitation toward biomedical care, as described above, others, especially from more modern Orthodox backgrounds, believed that medicine, created by God, was to be embraced and used. As one woman explained: *“I take an epidural. Why should I suffer? If the Almighty gave doctors such wisdom, why not use it?”* [36,p.144]. Another woman was quoted as saying: “u*sing technology is a statement of our interaction with God”* [40,p.275]. A few studies described “Dor Yeshorim”, a premarital genetic screening programme, widely used by Orthodox Jews worldwide for testing recessive genetic disorders, to prevent high-risk matches [37, 38, 40].

#### **Theme Two**: Holding Space or Holding Back: The Role of the Husband and the Community

##### a. Physical distance, spiritual support: the husband’s role in maternity

The Halacha (Jewish law) forbids men from watching the birth or having any physical contact with their spouses during this time. Women’s descriptions of this restriction were ambivalent; on the one hand, some women stated that their husbands provided spiritual support*: “He goes outside and reads some Psalms. He helps me from outside, holds my hand from there”* [36,p.147]. *“I give birth and my husband helps me spiritually. He can pray for me, and that is my biggest support”* [35,p.285]. On the other hand, a few women felt that the lack of support was too difficult. For example, following a pregnancy loss, a woman said: *“I needed someone to come and hug me…”* [42,p.350]. Despite the Halachic restriction, however, in Engelsman’s study, one woman was quoted saying: “*I do not let my husband move away from me at the birth itself*” [36,p.147].

##### b. The community: support, norms and taboos

The community is central in the lives of Orthodox Jewish people, offering support and solidarity. At the same time, expectations, norms, and taboos may shape and influence personal choices. Community support was described as especially vital during childbirth, where a female companion, often a family member, an experienced mother, or a doula from within the community, was present to provide support in the physical absence of the husband [34, 35, 38, 43].

However, in nearly all included studies, there were references, either directly or indirectly, to the community expectations and social pressures. In Engelsman et al.’s study, women described pressure to marry young and conceive soon after [36]. This was also described in Levitan et al.’s study: “*It’s a thing in our community, it’s a big thing: You get married –– you have a baby!”* [42,p.351]. Similarly, women spoke about the lack of support and the stigma surrounding pregnancy loss. One participant said, *“I definitely wasn’t telling friends…”* [42,p.351].

Prins-Engelsman and Cwikel explored the relationships between pregnancy and childbirth experiences, and postpartum depression (PPD) and general depression among Haredi women in Israel [44]. In their study, 22% of the women were at risk for PPD symptoms; this was higher in women from a Sepharadi sect (a more marginalised Orthodox group), and among women who were previously secular (therefore new to the community), indicating that lack of support may be a trigger.

#### **Theme Three**: Born to Become a Mother, Through Joy and Hardship

##### a. Reproduction as a religious obligation and identity

For Orthodox Jewish women, childbirth is not only a physical event, but a religious and Halachic calling, integral to their spiritual identity. Motherhood is seen as a Mitzvah (religious obligation) and an expression of feminine devotion. Even when involving pain or loss, many women interpret them as spiritual events that strengthen their connection to God and the community.

Women described the multidimensionality of pregnancy and childbirth; for them, it was not only a personal choice but a religious and social duty [34–36, 38, 40, 42, 43]. For example, one woman said: *“pregnancy for me is a God-given duty”* [38,p.1530]. Yet another woman stated that: “*This is our greatest influence on the world, that we bring children here and raise them and make them into good people…”* [43,p.776].

##### b. Hardship as a test of faith

Women described hardships and ordeals they were facing as a ‘test of faith’, as explained by one orthodox woman: “*A test of faith is when you’re given something that you would normally, not necessarily want. And your faith is tested…”* [39,p.73]. For Orthodox Jewish women, God has a plan, and He chooses women to carry out missions [38–41]. The more faith a woman has, the greater the challenge He will provide her [38, 39], and having a child with special needs is reserved for women with the greatest faith, as described by one woman: “i*t’s [*having a child with Down syndrome*] considered really a divine privilege [zchut]. You have to be on a very high level to embrace it that way”* [39,p.73].

Despite their religious beliefs, some women expressed their fear of facing these tests: *“Don’t give me such a task. I mean I don’t feel I could handle it’’* [39,p.73]. A few women told stories of special needs children who were given up for adoption or were institutionalised [38, 39]. Furthermore, the challenges of being pregnant were mentioned in all studies, with the first pregnancy perceived as the hardest, as one woman shared: “*I wasn’t prepared to get pregnant right away…the trouble was, well, this is the first time you’re together with a man…there’s just so much going on*” [43,p.780].

#### **Theme Fou**r: Praying Quietly: Coping Mechanisms

Women described prayers as a means that helped them cope with childbirth pain: *“The pain was strong and I felt in control. I was happy, because I was praying the whole time…”* [36,p.144].

Experiences of stillbirth and pregnancy loss were varied and personal. Some women struggled with isolation due to stigma within the community. For example, one participant shared, *“I was lonely, and I didn’t want to be alone”* [42,p.351]. For others, faith provided a sense of comfort and connection, as was reflected in another woman’s experience: *“it felt as though someone was holding me and I was not alone”* [41,p.927]

#### **Theme Five:** Keeping it Kosher: Religious and Maternity Care

In a few studies, the importance of adhering to Jewish law (Halacha) during pregnancy and childbirth was mentioned, including practices related to modesty, Shabbat observance, and family purity (Niddah) [34–36]. Engelsman and colleagues also described women’s ambivalent views when choosing the place of birth. While some preferred Orthodox hospitals that were familiar with their religious needs, others opted for secular hospitals. As one Israeli woman explained, *“Haredi hospitals don’t pay any special attention anymore to the sixth or seventh delivery… whereas if I have the baby at X Hospital [*secular*] I’ll be a queen, because they want special cases, and it’s also worth money”* [36,p.144].

However, Moreyossef and Chen found that among Orthodox Jewish women, the most significant factor contributing to birth satisfaction was not the physical environment, but rather the quality of interpersonal care. Kindness, empathy, and clear communication were stronger predictors of a positive birth experience than whether the setting was religiously appropriate [45].

## Discussion

This scoping review mapped the existing literature on the needs and experiences of Orthodox Jewish couples in maternity. Our primary finding is the limited literature on the subject. Although twelve relevant studies were identified, they covered a range of topics, and no study was conducted in the UK. Furthermore, most studies were conducted over a decade ago, with one study dating back to 1999. Contemporary research in the UK may yield different results, given the increasing integration of Orthodox Jewish women into British society and the workforce, as indicated by our Patient and Public Involvement and Engagement (PPIE) members.

One common thread across the identified studies is the integral role of spirituality in Judaism, particularly concerning childbirth, which was supported by the literature and comes as no surprise, given Judaism’s deeply rooted spiritual and cultural traditions [9, 14, 27, 46–48]. One participant explained: “*we take spirituality and we infuse it into every aspect of our daily lives*” [35,p.288]. Participants described the spirituality dimension as a means of supporting them through childbirth and hardships [34, 36, 38, 41].

Data extracted from our studies reveal a complex interplay between religious duties, community expectations and risk-based medicine. Religious beliefs were central in shaping women’s experiences and supporting decision-making around pregnancy, childbirth, and loss. Other papers emphasised the importance of adhering to the Halacha during pregnancy and childbirth, including modesty, Shabbat observance, kosher diet and intimacy restrictions between spouses [9, 11–13, 49].

Halachic laws guide all Orthodox Jews; however, the level of adherence and the interpretation of these ancient laws vary among Orthodox sects and rabbis. Modern Orthodox women tend to follow medical recommendations more closely, whereas Ultra-Orthodox women may regard rabbinical rulings as superior [12, 13]. The role of the rabbi in matters related to pregnancy and childbirth was demonstrated in the included studies and was more evident during times of uncertainty and complex decision-making, such as when invasive tests are needed after an abnormal screening result or for pregnancy spacing to support maternal emotional needs [34, 36–40, 43]. The rabbi was described as an authoritative and understanding figure [35, 36, 38–40, 43]. In her book *Conceiving Agency*, Raucher noted that contemporary rabbis often possess extensive knowledge beyond religious matters, enabling them to guide a wide range of topics, including health issues [18]. In the UK, a few rabbi-led medical advisory centres aim to bridge religion and modern medical practices by offering guidance and referral services to pregnant women [50, 51].

Rabbinical decisions can sometimes conflict with medical advice, leaving women confused as to which authority to follow. While some choose to follow medical recommendations, others will be supported by their rabbis and decline interventions. Gross and Shuval found that the main sources of resistance to risk-based medicine among Orthodox Jews are epistemological conflicts between faith and risk-based medicine, and concerns over perceived governmental control over private matters [37]. The first is based on the belief that prognosis, which is based on statistics and probabilities, is not reliable and cannot inform decision-making. Decisions during pregnancy should be based solely on diagnosis or certainties [37]. This idea was echoed in Teman et al.’s paper, where a woman stated: “*We are not going by probabilities and statistics […] you are looking at a different principle of what runs the universe”* [39,p.75]. The second rejection, according to Gross and Shuval, stems from the Orthodox community’s desire for autonomy. Accepting risk-based medicine and aligning with secular society, which only accepts “perfect babies” is considered to conform to modern values [37, 39]. Teman and colleagues argued that Orthodox women favour “religious certainty”, measured by trust in God and the understanding that He is in control, over “secular certainty”, based on probabilities [39]. However, Raucher suggested that, drawing from their embodied experiences, Haredi women develop reproductive autonomy within a patriarchal framework. With pregnancy often kept private, rejecting medical advice can serve as a subtle way to assert control over their body and life [18].

A central Halachic concept is Shabbat observance. The challenges it presents to couples during labour prompted a UK rabbi, Mr. Jacobson, to draft the “*Childbirth in Shabbos: a practical guideline*”, in half-Hebrew half-English [49]. This document was shared with us and highlights the challenges of childbirth on Shabbat, during which Orthodox Jews are prohibited from using a phone, electric devices, driving a car, or writing. Unless the situation is deemed life-threatening, the couple may face restrictions on using certain hospital facilities, such as electric doors and sensor-activated faucets and toilets [49]. Rabbi Jacobson reminds couples that “*Shabbat is an everlasting bond between Hashem* [God] *and the Jewish nation, and therefore its sanctity is very great”* [49,p.3].

Although not discussed in length in the included studies, Pikuah Nefesh, the preservation of human life that takes precedent over almost any other law, is a core concept in Judaism [47, 52]. Rabbi Jacobson referred to labouring on Shabbat and instructed: *“…an ישראל* [Israeli] lif*e takes precedence over the Shabbat…”* [49,p.3], suggesting that a labouring woman is exempt from observing the Shabbat. Another example of Pikuah Nefesh is where a foetus presents a risk to the mother; in this case, there will usually be a greater tendency towards additional pregnancy tests or interventions, and in some cases, the rabbi will permit abortion [52]. In contradiction to this concept, a few scholars found that some women conceived faith and religious obligation as the highest commitment and were willing to risk their lives and continue their high-risk pregnancy or go through five or more caesarean sections to fulfil their duty [18, 43].

Shabbat and holy days restrictions, coupled with the law of Niddah, may lead women in labour and the immediate postpartum period to feel isolated and lonely. Niddah prevents the husband from having any physical contact with his wife from the mark of active labour or any vaginal bleeding for a few days or weeks after birth [27]. Since interpretation of the Jewish law may vary slightly, depending on the sect and personal perceptions, some husbands stay in the room during childbirth, while standing behind a curtain, while others leave the room entirely. Semenic found that 30% of Orthodox women had no support during childbirth, and only 37% of the husbands remained in the room [34].

The rationale of the law of Niddah and the complexity it creates around sexuality and fertility are beyond the scope of this paper. However, the implications for birth were discussed in a few of the included studies and portray a mixed picture, with some women describing the spiritual support the husband provides, without physical contact, as powerful and sufficient. In contrast, others described the challenges it creates. In her case study on postpartum psychosis (PPP), Posmontier described how the absence of the husband in the days following birth, due to Jewish holy days, combined with sleep deprivation, led to PPP [53]. Following birth, the new mother was left alone after her husband had left the hospital to observe a Jewish holy day, during which phone use is also prohibited. Recalling the experience, the woman stated: *“secluded from the whole world, since I couldn’t speak to anyone… I just felt very lonely, very homesick. I felt like no one was really paying attention to me”* [53,p.171]. In the days that followed, she started exhibiting symptoms of psychosis that eventually led to a full psychotic episode [53].

This story is a documentation of a rare case that had multiple triggers; however, it highlights the challenges of newly Orthodox mothers who may find themselves struggling to cope alone and may not reach out for help due to stigma and taboo. This was illustrated in Levitan’s study, where a woman who felt that taboos around pregnancy loss contributed to feelings of loneliness. She stated: *“I would like to talk about it. It would have been healing for me to talk about it”* [42,p.351].

The lack of physical support from the husband during Niddah has led to well-established community doula services available in many countries, including the UK [9, 19, 40, 43]. In his book, describing the Orthodox community in Manchester and their maternity experiences, Kasstan wrote: *“frum* [Orthodox] *doulas claim to be advantageous for the local NHS authority because they can contribute to making mainstream maternity services more accessible for Haredi Jews”* [19,p.146]. In our included studies, women described the importance of the community in their lives and the support it offers from pregnancy, through the postpartum period, and beyond [34–36, 38, 43]. Furthermore, Hatzola, an Orthodox-run ambulance service, was highlighted by both the literature and PPIE members as a valued, culturally sensitive resource, though concerns were raised by PPIE members regarding privacy due to it being staffed by community members [19, 49]. Another Haredi UK-based charity is Interlink, which brings together Haredi community organisations involved in maternity care, as well as statutory providers and commissioners. The organisation collaborates with Homerton maternity services and several community charities and aims to improve services and communication for Haredi residents in Hackney [54]. A variety of other local services were also shared with us by PPIE members, including free (*Gmach*) and private options. This richness reflects the community’s need for culturally tailored support.

Although we did not identify any maternity-specific studies from the UK, studies of wider healthcare experience have revealed a disconnect between Orthodox families and healthcare professionals. Women felt that healthcare staff lacked cultural understanding, while professionals viewed the community as self-sufficient and believed their services were unnecessary [17, 55]. One nurse remarked, *“It’s like a community who don’t need outsiders”* [55,p.82]. Women mentioned in the literature preferred seeking advice from experienced community members rather than healthcare professionals, who are sometimes seen as lacking both cultural awareness and personal experience. For example, in Kasstan’s book, one woman described Orthodox doulas as providing continuity of care, a model she felt no longer exists within the NHS [19]. Similarly, in Abbott’s study, “Lay and Professional Views on Health Visiting in an Orthodox Jewish Community”, a community stakeholder questioned the credibility of health visitors, asking: *“How can you regard as a professional breastfeeding expert someone who’s not had a child themselves? And they don’t even know the mechanics of breastfeeding”* [55,p.83].

Relying on the community can be rooted in the lack of trust in the health system, found in a few studies [38–40]. Local reports supported this finding, indicating that in some cases, a lack of trust led to poor adherence to public health recommendations [15–17, 20, 22, 56]. On the positive side, lower acceptance of medical intervention among Orthodox Jewish women has been linked to significantly lower Caesarean section (CS) rates of 8.56% in Israel and 3% in Manchester, UK, compared to the general population’s reported rate of 14.8% and 26% in Israel and the UK, respectively [19, 23]. CS are correlated with complications for both mothers and babies, including increased mortality, and are therefore only recommended in cases with a clear medical indication [57]. Nevertheless, a recent Cambridge publication indicates that 3% and 4% of CS in the UK are performed for maternal request and previous CS, respectively [57]. Therefore, by preferring vaginal birth, Orthodox women may reduce their risk for CS related complications.

Despite the many benefits of its support, the intensity of the community was portrayed in the included studies as a double-edged sword. Social expectations, taboos and Halachic restrictions led some women to feel isolated and were sometimes seen as having a greater influence on decision-making, rather than personal choice [36, 38–44]. Other studies found that the social pressure to conceive and taboos around loss and mental health in Orthodox Jewish communities are inhibiting factors preventing Orthodox women from seeking medical and psychological help [15, 16, 26, 58, 59].

## Strengths and limitations

The diverse expertise of the research team members is the key strength in our review. Our team included a midwife, a professor of health and social care, and a professor of maternal and child health, each contributing a unique perspective that enriched the analysis. Additionally, the first author is an Israeli living in the UK, bringing a bicultural understanding that supported the interpretation of the data. While not from an Orthodox background, familiarity with Orthodox practices informed the interpretation of the findings. Three Orthodox women from our PPIE group, including two midwives, reviewed the paper draft, and their thoughtful feedback helped strengthen the published version.

We were also systematic in our search strategy and analysis, following a structured scoping review methodology to ensure transparency and allow for future replication. This included a thorough search of pre-agreed databases, careful screening and selection of studies, and an iterative analytical process. All decisions were made as a team.

This review has several limitations, with the main one being the absence of UK-based studies in our synthesis. As a result, we were unable to answer the review sub-questions, and the applicability of findings to the UK context may be limited. Experiences in countries like Israel, where Orthodox Jewish communities are more numerous and have access to culturally sensitive services, may differ significantly.

The literature concerning the needs and experiences of Orthodox Jewish couples in maternity care is limited, and we had to make adaptations to our search strategy. Most of the included studies were qualitative, though two were quantitative and required conversion to be analysed thematically, potentially limiting nuance. One study had a small sample size (three participants), reducing generalisability. Half of the included studies were conducted over a decade ago and may no longer reflect the current realities. Additionally, some of the included studies did not discuss our topic of inquiry directly, and we had to read and interpret them carefully to extract relevant data. Furthermore, while we aimed to understand the interaction of Orthodox couples with maternity services, no study investigated male partners.

A few of the included papers were based on the same underlying datasets; however, we chose to include them as separate studies, as they explored different aspects and presented quotations that contributed meaningfully to our analysis. While literature in Hebrew may exist, time constraints did not allow for a broader search. Furthermore, the cyber-attack on the British Library at the time the review was undertaken prevented access to academic theses.

Finally, as we relied on secondary data, we were limited to the quotes and interpretations provided by the original authors of the papers. This limited our ability to explore the original data in full, potentially identifying commonalities among women whose perspectives differed from the main narratives.

## Conclusions

This scoping review highlights the limited literature on the needs and experiences of Orthodox Jewish couples in maternity care, with a significant gap regarding the experiences of male partners. Although twelve relevant studies were identified, none were conducted in the UK, and half were conducted over a decade ago, limiting their relevance to the current UK context.

The findings highlight the important role of religion and community in shaping women’s experiences and decision-making during pregnancy, childbirth, and loss. While the community provides significant support, it can also impose expectations and restrictions.

There is a clear need for high-quality, contemporary research in the UK to explore the needs of Orthodox Jewish couples and to inform equitable and culturally sensitive maternity services.

## Contributors

**Michal Rosie Meroz:** conceptualisation (lead); search (lead); writing the original draft (lead); analysing (lead); writing the review (lead); editing (equal); guarantor. **Carol Rivas:** conceptualisation (supporting); search (supporting); analysis (supporting); writing the original draft (supporting); editing (equal). **Christine McCourt:** conceptualisation (supporting); analysis (supporting); writing the original draft (supporting); editing (equal).

## Data Availability

This is a review and the full review tables and extraction sheets can be made available if required

## Acknowledgment

We would like to express our deepest appreciation to Sara Neiman and Sara Barnett from London for reading our draft and providing their valuable insights.

Many thanks to Tova Schprecher, the project’s PPIE lead from Manchester, for all her support throughout.

## References list

1. Graham D, Boyd J. Jews in the UK today: key findings from the JPR national Jewish identity survey. 2024 [cited 2025 Jan 27]. Available from: https://www.jpr.org.uk/reports/jews-uk-today-key-findings-jpr-national-jewish-identity-survey

2. NHS Race Health Observatory. Review of NHS health communications with (and for) the Jewish community. 2024 [cited 2024 Feb 17]. Available from: https://nhsrho.org/research/health-communications-report-and-resources-to-improve-access-to-nhs-services-for-jewish-communities/

3. Staetsky D. Haredi Jews around the world: population trends and estimates. Institute for Jewish Health Policy (JPR). 2022 [cited 2025 Jan 27]. Available from: https://www.jpr.org.uk/reports/haredi-jews-around-world-population-trends-and-estimates

4. Staetsky LD, Boyd J. Strictly Orthodox rising: what the demography of British Jews tells us about the future of the community. 2015 [cited 2025 Jan 27]. Available from: https://archive.jpr.org.uk/object-uk285#:∼:text=In20particular2C20we20highlight20how,its20below20replacement20level%20fertility.

5. Flint-Ashery S. Spatial behaviour in Haredi Jewish communities in Great Britain. 1st 2020. ed. Cham: Springer International Publishing; 2020.

6. Limb M. Fertility rate in England and Wales fell to a record low in 2023. BMJ (Online). 2024;387:q2397. doi: 10.1136/bmj.q2397

7. Simhi M, Yoselis A, Sarid O, Cwikel J. Hidden figures: are Ultra-Orthodox Jewish women really so different when it comes to health care? J Relig Health. 2020;59(3):1398–420. doi: 10.1007/s10943-019-00862-2

8. Bash DM. Jewish religious practices related to childbearing. J Nurse. 1980;25(5):39–42. doi: 10.1016/0091-2182(80)90162-7

9. Berkowitz B. Cultural aspects in the care of the orthodox Jewish woman. J Mid Womens Health. 2008;53(1):62–7. doi: 10.1016/j.jmwh.2007.07.008.

10. De Sevo MR. Jewish traditions in pregnancy & childbirth. Nurs Women’s Health. 1997;1(4):46–9. doi: 10.1111/nuf.12320

11. Cassar L. Cultural expectations of Muslims and Orthodox Jews in regard to pregnancy and the postpartum period: a study in comparison and contrast. Int J Childbirth Educ. 2006;21(2):27–41 [cited 2025 Mar 12]. Available from: https://research.ebsco.com/c/v5rb7r/viewer/pdf/6g3xyoemo5?route=details

12. Lewis JA. Jewish perspectives on pregnancy and childbearing. MCN Am J Matern Child Nurs. 2003;28(5):306–12 [cited 2025 Mar 12]. Available from: https://oce.ovid.com/article/00005721-200309000-00008/HTML

13. Noble A, Rom M, Newsome-Wicks M, Engelhardt K, Woloski-Wruble A. Jewish laws, customs, and practice in labor, delivery, and postpartum care. J Transcult Nurs. 2009;20(3):323–33. doi: 10.1177/104365960933

14. Bodo K, Gibson N. Childbirth customs in Orthodox Jewish traditions. Can Fam Physician. 1999 Mar;45:682–6. PMID: 10099807

15. Hackney and City. Health needs assessment: Orthodox Jewish community in Stamford Hill North Hackney. 2018.

16. Wineberg J, Mann S. Salford Jewish community health research report 2015. 2016 [cited 2025 Feb 06]. Available from: https://archive.jpr.org.uk/object-uk370

17. Phillips N, Hakak Y. Navigating Ultra-Orthodox Jewish motherhood in the United Kingdom. JourMS. 2023 [cited 2025 Feb 06]. Available from: https://jourms.org/navigating-ultra-orthodox-jewish-motherhood-in-the-united-kingdom-the-perspectives-on-the-understanding-and-challenges-of-social-work-support-through-the-haredi-mothers-lens/

18. Raucher M. Conceiving agency: reproductive authority among Haredi women. 1st ed. Bloomington, Indiana: Indiana University Press; 2020.

19. Kasstan B. Making bodies kosher: the politics of reproduction among Haredi Jews in England. First ed. New York: Berghahn Books; 2019.

20. Letley L, Rew V, Ahmed R, Habersaat KB, Paterson P, Chantler T, et al. Tailoring immunisation programmes: using behavioural insights to identify barriers and enablers to childhood immunisations in a Jewish community in London, UK. Vaccine. 2018;36(31):4687–92. doi: 10.1016/j.vaccine.2018.06.028

21. Spitzer J. A guide to the Orthodox Jewish way of life for healthcare professionals. Sixth ed. London: Dr J Spitzer; 2019.

22. Interlink. Community insight report: how Charedi children and young people access child health and development services. Hackney: Healthwatch Hackney City and Hackney Clinical Commissioning Group; 2014.

23. Arbel Y, Bar-El R. Cesarean sections and family planning among Ultra-Orthodox Israeli Jews. J Relig Health. 2024;63(4):2599–632. doi: 10.1007/s10943-024-02026-3

24. Mayhew L, Harper G, Waples S. Counting Hackney’s population using administrative data: an analysis of change between 2007 and 2011. Report commissioned by Hackney Council, Mayhew Harper Associates, London. 2011.

25. Graham D, Waterman S. Underenumeration of the Jewish population in the UK 2001 Census. Population space and place. 2005;11(2):89–102. doi:10.1002/psp.362

26. Zauderer C. Maternity care for Orthodox Jewish Couples: Implications for Nurses in the Obstetric Setting. Nurs Womens Health. 2009;13(2):112–20. doi: 10.1111/j.1751-486x.2009.01402.x

27. Feldman P. Sexuality, birth control and childbirth in Orthodox Jewish tradition. CMAJ. 1992;146(1):29–33. PMID: 1728349

28. Lutwak RA, Ney AM, White JE. Maternity nursing and Jewish law. MCN Am J Matern Child Nurs. 1988;13(1):44–6. [cited 2024 Dec 05]. Available from: https://oce.ovid.com/article/00005721-198801000-00014/HTML

29. Grimshaw J. A knowledge synthesis chapter. In: Canada: Canadian Institutes of Health Research.

30. Arksey H, O’Malley L. Scoping Studies: towards a methodological framework. Int. J. Soc. Res. Methodol. 2005;8(1):19–32. doi: 10.1080/1364557032000119616

31. Moher D, Liberati A, Tetzlaff J, Altman DG, the PRISMA Group. Reprint— preferred reporting items for systematic reviews and meta-analyses: The PRISMA Statement. Phys Ther. 2009;89(9):873–80. doi:10.1093/ptj/89.9.873 pmid:19723669

32. Peters MDJ, Marnie C, Tricco AC, Pollock D, Munn Z, Alexander L, et al. Updated methodological guidance for the conduct of scoping reviews. JBI Evid Synth. 2020;18(10):2119–26. doi: 10.11124/JBIES-20-00167

33. Thomas J, Harden A. Methods for the thematic synthesis of qualitative research in Systematic Reviews. BMC Med Res Methodol. 2008;8(1):45. doi: 10.1186/1471-2288-8-45

34. Semenic SE, Callister LC, Feldman P. Giving birth: the voices of Orthodox Jewish women living in Canada. J Obstet Gynecol Neonatal Nurs. 2004;33(1):80–7. doi: 10.1177/0884217503258352

35. Callister LC, Semenic S, Foster JC. Cultural and Spiritual Meanings of Childbirth: Orthodox Jewish and Mormon Women. j holist nurs. 1999;17(3):280–95. doi: 10.1177/089801019901700305

36. Engelsman SP, Huss E, Cwikel J. How Ultra-Orthodox (Haredi) Israeli women cope with normative and difficult pregnancy and childbirth experiences. Nashim. 2018(33):136–57. doi: 10.2979/nashim.33.1.07.

37. Gross SE, Shuval JT. On knowing and believing: prenatal genetic screening and resistance to ‘risk-medicine’. Health Risk Soc. 2008;10(6):549–64. doi: 10.1080/13698570802533721

38. Ivry T, Teman E, Frumkin A. God-sent ordeals and their discontents: Ultra-Orthodox Jewish women negotiate prenatal testing. soc sci med. 2011;72(9):1527–33. doi: 10.1016/j.socscimed.2011.03.007

39. Teman E, Ivry T, Bernhardt BA. Pregnancy as a proclamation of faith: Ultra-Orthodox Jewish women navigating the uncertainty of pregnancy and prenatal diagnosis. Am J Med Genet A. 2010;155A(1):69–80. doi: 10.1002/ajmg.a.33774

40. Teman E, Ivry T, Goren H. Obligatory Effort [Hishtadlut] as an explanatory model: a critique of reproductive choice and control. Cult Med Psychiatry. 2016;40(2):268–88. doi: 10.1007/s11013-016-9488-5

41. Hamama-Raz Y, Hartman H, Buchbinder E. Coping with Stillbirth among Ultraorthodox Jewish Women. Qual Health Res. 2014;24(7):923–32. doi: 10.1177/1049732314539

42. Levitan F, Frei-Landau R, Sabar-Ben-Yehoshua N. “I just needed a hug”: culturally-based disenfranchised grief of Jewish Ultraorthodox women following pregnancy loss. Omega (Westport). 2025;91(1):342–60. doi: 10.1177/00302228221133864

43. Teman E, Ivry T. Pregnancy and the reproductive Habitus of Ultra-Orthodox Jewish women. Med Anthropol. 2021;40(8):772–84. doi: 10.1080/01459740.2021.1961244

44. Prins-Engelsman S, Cwikel J. Postpartum depression symptoms in various Sects of Ultra-Orthodox society in Israel. J Relig Health. 2023;62(5):3327–46. doi: 10.1007/s10943-023-01745-3

45. Moryossef IG, Chen KO. Predicting maternal satisfaction with delivery services among Ultra-Orthodox Jewish mothers in public hospitals. Int J Nurs Educ. 2021;13(3):36–43 [cited 2025 Mar 30]. Available from: https://web.p.ebscohost.com/ehost/pdfviewer/pdfviewer?vid=0&sid=4baa8536-22db-4cfb-83f8-e8c9496b7569%40redis

46. Jacob A, Goldzweig G, Dar A, Igra L, Hasson-Ohayon I. Public stigma toward women, mothers and non-mothers, with serious mental illness in Jewish Ultraorthodox society. Ment Health Relig Cult. 2024;27(1):27–43. doi: 10.1080/13674676.2024.2321626

47. Gabbay E, McCarthy MW, Fins JJ. The care of the Ultra-Orthodox Jewish patient. J Relig Health. 2017;56(2):545–60. doi: 10.1007/s10943-017-0356-6

48. Nov-Klaiman T, Raz AE, Hashiloni-Dolev Y. A test of faith? attitudes of Ultraorthodox Jewish parents of children with Down Syndrome toward prenatal testing. Disability & society. 2024;39(1):192–212. doi: 10.1080/09687599.2022.2070059

49. Jacobson, A. Childbirth on shabbos: a practical guide. 2009.

50. MARS: Meidcal Advocacy & Referral Service [Internet]. [cited 2025 Apr 07]. Available from: https://marsorg.com/

51. Tahareinu: Transforming Lives with Medical Solutions [Internet]. [cited 2025 Apr 07]. Available from: https://tahareinu.com/

52. Schenker JG. The beginning of human life: status of embryo. Perspectives in Halakha (Jewish Religious Law). J Assist Reprod Genet. 2008;25(6):271–6. doi: 10.1007/s10815-008-9221-6

53. Posmontier B, Fisher KM. A narratology of postpartum psychosis in an Orthodox Jewish woman. Perspect Psychiatr Care. 2014;50(3):167–77. doi: 10.1111/ppc.12037

54. Interlink [Internet]. [cited 2025 Mar 31]. Available from: https://www.interlink-foundation.org.uk/upcoming-trainings/

55. Abbott S. Lay and professional views on health visiting in an orthodox Jewish community. Br J Community Nurs. 2004;9(2):80–6. doi: 10.12968/bjcn.2004.9.2.12424

56. JuMP. Mothers health support project: Pilot report, October 2015-March 2016. HJMT, Interlink, Beis Brucha. 2016.

57. Dahlke JD, Chauhan SP. Caesarean section delivery. Cambridge: Cambridge University Press; 2025 [cited 2025 July 22]. Available from: https://www.cambridge.org/core/elements/caesarean-section-delivery/443C3305B3EE207736564D61C5722B90

58. Bina R. Seeking help for postpartum depression in the Israeli Jewish Orthodox community: factors associated with use of professional and informal help. Women Health. 2014;54(5):455–73. doi: 10.1080/03630242.2014.897675

59. Bina R, Glasser S. Factors associated with attitudes toward seeking mental health treatment postpartum. Women Health. 2019;59(1):1–12. doi: 10.1080/03630242.2017.1421286

